# Age modulates protein-outcome associations in deceased donor kidneys

**DOI:** 10.1101/2023.03.31.23288011

**Authors:** Philip D Charles, Rebecca H Vaughan, Sarah Fawaz, Simon Davis, Priyanka Joshi, Iolanda Vendrell, Roman Fischer, Benedikt M Kessler, Edward J Sharples, Alberto Santos, Rutger J Ploeg, Maria Kaisar

## Abstract

**Background:** Organ availability limits kidney transplantation, the best treatment for end-stage kidney disease. Globally, deceased donor acceptance criteria have been relaxed to include older donors, which comes with a higher risk of inferior posttransplant outcomes. Donor age, although negatively impacts transplant outcomes, lacks granularity in predicting graft dysfunction. Better donor kidney assessment and characterization of the biological mechanisms underlying age-associated donor organ damage and transplant outcomes is key to improving donor kidney utilisation and transplant longevity.

**Methods:** 185 deceased pretransplant biopsies (from brain and circulatory death donors aged 18-78 years) were obtained from the Quality in Organ Donation (QUOD) biobank and proteomic profiles were acquired by mass spectrometry. Machine learning exploration using prediction rule ensembles guided LASSO regression modeling of kidney proteomes that identified protein signatures and biological mechanisms associated with 12-m posttransplant outcome. Data modeling was validated on held-out data and contextualised against published spatially resolved kidney injury related transcriptomes.

**Results:** Our analysis highlighted that outcomes were best modeled using combination of donor age and protein abundance signatures, revealing 539 proteins with these characteristics. Modeled age:protein interactions demonstrated stronger associations with transplant outcomes than age and protein alone and revealed mechanisms of kidney injury including metabolic changes and innate immune responses correlated with poor outcome. Comparison to single-cell transcriptome data suggests protein-outcome associations to specific cell types.

**Conclusions:** Molecular signatures resulted from integration of donor age and proteomic profiles in deceased donor kidney biopsies offer the potential to develop improved pretransplant organ assessment and aid decisions on perfusion interventions.

## INTRODUCTION

Kidney transplantation is the optimal treatment for end-stage kidney disease (ESKD). Compared to dialysis, transplantation increases life-expectancy, improves quality of life and is cost-effective. Limited availability of suitable donor kidneys impedes treatment, and often prolongs dialysis, increasing morbidity and mortality. Deceased donor organ shortages, living donation decline in some countries, and ageing populations drive increased utilization of older deceased donor kidneys, now comprising more than half of offered organs ^1,2^.

Ageing correlates with decline of organ function, evidenced in kidneys by nephron loss and histological lesions, such as tubular atrophy, interstitial fibrosis, glomerulosclerosis, and arteriosclerosis. Increasing age links with declining function as kidneys have fewer functioning glomeruli, reduced renal mass, podocyte dysfunction, and impaired cellular repair ^3^. Glomerular diseases are also more common and associated with worse disease outcomes in older patients ^4^.

Age correlates with increased prevalence of Chronic Kidney Disease (CKD) and accelerated transition to chronic disease from Acute Kidney Injury (AKI) ^5^. In organ donation and transplantation, older donors are more likely to have suffered from additional comorbidities such as diabetes, hypertension or cardiovascular disease, and organs from these ‘higher risk’ donors have higher rates of graft dysfunction or loss ^6^. As donor age is a major factor in determining transplant outcomes, is incorporated in clinical scoring algorithms to guide kidney allocation decisions ^7,8^. Current front-line indices that also include clinical factors such as terminal serum creatinine, history of hypertension and diabetes ^8,9^ show consistent performance across demographics but lack granular accuracy for donor kidney quality assessment ^10^, resulting in increased organ decline of potentially viable organs and uncertainty about transplant longevity ^11^.

Molecular analyses of biopsies from donor kidneys offer higher resolution assessment of organ state, but robust associations with transplant outcomes are yet to be established. Deceased donors are frequently assessed as having sustained kidney injury (i.e. AKI) based on serum creatinine levels ^12^, however this metric alone is a poor indicator of long term outcomes ^12–15^. Transcriptomic and proteomic studies of donor kidney biopsies provide molecular granularity, highlighting the role of metabolic changes and inflammatory response pathways ^16,17^, but limited availability of well curated and annotated clinical samples restricts investigations ^18^. Integration of molecular phenotypes using machine learning approaches from analysis of donor kidneys with clinical and demographic factors may enable prognostic judgements about kidney donation, taking the next steps towards precision medicine ^19^.

Age-associated changes to the donor kidney transcriptome and proteome modify the organ capacity to recover from ischemia-reperfusion injury and may influence suboptimal outcomes in graft function ^17^. However, there is limited data on whether ageing changes how proteomic profiles in donor kidneys associate with transplant outcome. Our study seeks to disentangle the effects of ageing on kidney proteome alterations and define associations with transplant outcome while in parallel provides key insights to the key biological processes that may implicated in outcome-relevant changes. Overall our investigation has the potential to lead to development of improved donor kidney assessment and novel intervention targets for organ reconditioning.

## METHODS

### Study Design

Deceased donor pretransplant biopsies (n=186; 1 sample excluded during data processing) were obtained from the Quality in Organ Donation (QUOD) biobank, a national multi-center UK wide bioresource of deceased donor clinical samples acquired during donor management and organ procurement. Pretransplant biopsies were obtained from Donation after Brain Death (DBD) donors and Donation after Circulatory Death (DCD) donors during back table preparation.

Kidneys were selected to cover the outcome continuum i.e. the range of estimated Glomerular Filtration Rate (eGFR) in the recipient at 12 months posttransplant (henceforth, ‘eGFR12’), from primary non-function to eGFR12>80 ml/min/1.73 m^2^ from donors aged 18 to 78 years. To minimize the impact of recipient factors, we only included kidneys for which the contralateral kidney was transplanted with similar outcome, both kidneys having eGFR12< 30 ml/min/1.73 m^2^, between 30 and 60 ml/min/1.73 m^2^, or > 60 ml/min/1.73 m^2^). Samples were linked to corresponding donor and recipient demographic and clinical metadata, provided by NHS Blood and Transplant National Registry.

### Study Approval and Ethics statement

Informed consent from donor families was obtained prior to sample procurement consistent with the Declaration of Helsinki. Collection of QUOD samples and research ethics approval was provided by QUOD (NW/18/0187). The clinical and research activities being reported are consistent with the Principles of the Declaration of Istanbul as outlined in the ‘Declaration of Istanbul on Organ Trafficking and Transplant Tourism’.

### Sample Analysis and Statistical Analyses

Please see Supplementary Methods.

## RESULTS

### Associations of Clinical Variables with Transplant Outcome are Dominated by Donor Age

The selected cohorts were representative of the deceased donor population when demographic and clinical characteristics were considered (Table 1). All 51 donor variables used in our modeling are listed in Supplementary Table ST1. eGFR12 was strongly inversely correlated with donor age (R^2^=0.2107; p=5.0×10-11; Supplementary Table ST1). UK Kidney Donor Risk Index (R^2^ =0.2042; p=2.5×10-9; Supplementary Table ST1) explained less variance in eGFR12 than donor age alone, despite representing a synthesized risk prediction of poor outcomes from UK deceased donors.

**Table 1:** Donor and recipient clinical and demographic variables. Donor kidney associated metadata. Samples are subdivided by donor type for information purposes. Numerical variables are given as mean ± standard deviation. Categorial variables are given as frequency alongside percentage of total cohort. Correlation values and associated p values given are for the correlation between each variable and Recipient eGFR at 12 months posttransplant (i.e. not a comparison between DBD and DCD)

|  | DBD | DCD | Total | Proportion of<br>Variance<br>Explained | Association<br>p-value |
| --- | --- | --- | --- | --- | --- |
| n | 100 | 85 | 185 |  |  |
| Recipient kidney function (mean<br>eGFR, mL/min per 1.73 m<sup>2</sup>)<br>at 12 Months Posttransplant<br>(eGFR12) | 53.47<br>± 28.29 | 47.38<br>± 25.17 | 50.67<br>± 27.00 | - | - |
| Recipient kidney function (mean<br>eGFR, mL/min per 1.73 m<sup>2</sup>)<br>at 3 Months Posttransplant | 54.26<br>± 28.94 | 48.14<br>± 21.20 | 51.47<br>± 25.81 | 0.6589 | 6.7079×10<sup>-44</sup> (***) |
| Donor Age, y | 48.37<br>± 14.98 | 49.60<br>± 12.81 | 48.94<br>± 14.00 | 0.2107 | 5.0067×10<sup>-11</sup> (***) |
| Donor Sex<br>Female<br>Male | 50 (50.0%)<br>50 (50.0%) | 31 (36.5%)<br>54 (63.5%) | 81 (43.8%)<br>104 (56.2%) | 0.0051 | 3.3568×10<sup>-1</sup> |
| Donor Ethnicity<br>White<br>Other | 96 (96.0%)<br>4 (4.0%) | 83 (97.6%)<br>2 (2.4%) | 179 (96.8%)<br>6 (3.2%) | 0.008 | 9.1804×10<sup>-1</sup> |
| Donor Weight, kg | 80.26<br>± 17.97 | 80.60<br>± 15.34 | 80.42<br>± 16.77 | 0.0017 | 5.7320×10<sup>-1</sup> |
| Donor Height, cm | 171.19<br>± 9.96 | 171.95<br>± 8.99 | 171.54<br>± 9.51 | 0.0229 | 3.9808×10<sup>-2</sup> (*) |
| Donor S-Cr terminal, umol/l | 86.76<br>± 54.60 | 68.14<br>± 28.14 | 78.04<br>± 45.08 | 0.0055 | 3.3332×10<sup>-1</sup> |
| Donor Cold Ischemia Time, h | 14.70<br>± 4.35 | 13.92<br>± 5.58 | 14.34<br>± 4.95 | 0.0002 | 8.3482×10<sup>-1</sup> |
| Donor Cause of death<br>Trauma<br>Other | 7 (7.0%)<br>93 (93.0%) | 11 (12.9%)<br>74 (87.1%) | 18 (9.7%)<br>167 (90.3%) | 0.0396 | 6.5924×10<sup>-3</sup> (*) |
| Donor UKKDRI | 1.11<br>± 0.47 | 1.08<br>± 0.45 | 1.10<br>± 0.46 | 0.2042 | 2.5337e-09 (***) |
| Donor Delayed Graft Function<br>Immediate<br>Delayed<br>Primary Non-Function<br>Not Reported | 73 (73.0%)<br>17 (17.0%)<br>0 (0.0%)<br>10 (10.0%) | 54 (63.5%)<br>18 (21.2%)<br>1 (1.2%)<br>12 (14.1%) | 127 (68.6%)<br>35 (18.9%)<br>1 (0.5%)<br>22 (11.9%) | 0.025 | 1.3182e-01 |
| Recipient Age, y | 49.25<br>± 14.36 | 51.80<br>± 12.68 | 50.42<br>± 13.64 | 0.1117 | 3.3146e-06 (***) |
| Recipient Sex<br>Female<br>Male | 46 (46.0%)<br>54 (54.0%) | 21 (24.7%)<br>64 (75.3%) | 67 (36.2%)<br>118 (63.8%) | 0.0005 | 7.5185e-01 |
| Recipient Ethnicity<br>White<br>Other | 72 (72.0%)<br>28 (28.0%) | 65 (76.5%)<br>20 (23.5%) | 137 (74.1%)<br>48 (25.9%) | 0.0191 | 7.4663e-01 |
| HLA Mismatch Groups<br>0 Mismatches<br>0 DR and 0/1 B<br>0 DR and 2 B<br>or 1 DR and 0/1 B | 16 (16.0%)<br>30 (30.0%)<br>44 (44.0%)<br>10 (10.0%) | 2 (2.4%)<br>24 (28.2%)<br>49 (57.6%)<br>10 (11.8%) | 18 (9.7%)<br>54 (29.2%)<br>93 (50.3%)<br>20 (10.8%) | 0.03 | 1.3682e-01 |
| Donor/Recipient Sex Mismatch<br>No<br>Yes | 58 (58.0%)<br>42 (42.0%) | 26 (30.6%)<br>59 (69.4%) | 84 (45.4%)<br>101 (54.6%) | 0.0017 | 5.7505e-01 |

### Regression Modeling of Donor Kidney Proteomes and Clinical Metadata Identifies Outcome-Associated Proteins

Proteomic analysis of biopsies from donor kidneys quantified 2984 proteins with 50% or less missing values (out of 7790 identified protein groups in total; Supplementary Figures SF1A and SF2) over 185 samples and 20 interspersed sample pools as internal controls. Analysis of sample pools showed minimal mass spectrometry-related variance (squared mean pairwise Z-corrected Pearson’s r=0.94).

To assess individual protein relationships with outcome, we adopted a descriptive modeling approach, using 2/3 subset (‘discovery set’; n=118) for data exploration and modeling, retaining a held-out 1/3 subset (‘evaluation set’; n=61) to check our results generalized to unseen data, selected by stratified random sampling across eGFR12 tertiles.

Six of the donor kidney biopsies analyzed were paired (3 pairs from 3 donors); these were analyzed separately from the discovery and evaluation sets to assess intra-donor reproducibility. All three pairs showed high correlation of protein intensity values between donor pairs (Pearson’s r=0.71, 0.92 and 0.91; Supplementary Figure SF1B).

For an unbiased assessment of key relationships between clinical variables, protein measurements, and outcome, we used iterative Prediction Rule Ensemble ^20^ (PRE) learning on our training set to select features among the set of quantified proteins and donor type-independent clinical variables available pretransplant (Figure 1, inset). PRE modeling uses regularization to generate a minimal model (which prevents overfitting). To investigate the multitude of ways in which clinical factors might interact with protein associations with outcome we expanded the space of proteins considered for interaction with clinical variables by repeating this modeling 2000 times, excluding proteins identified in the rule ensemble at each iteration from future iterations while retaining all clinical variables.

**Figure 1:**
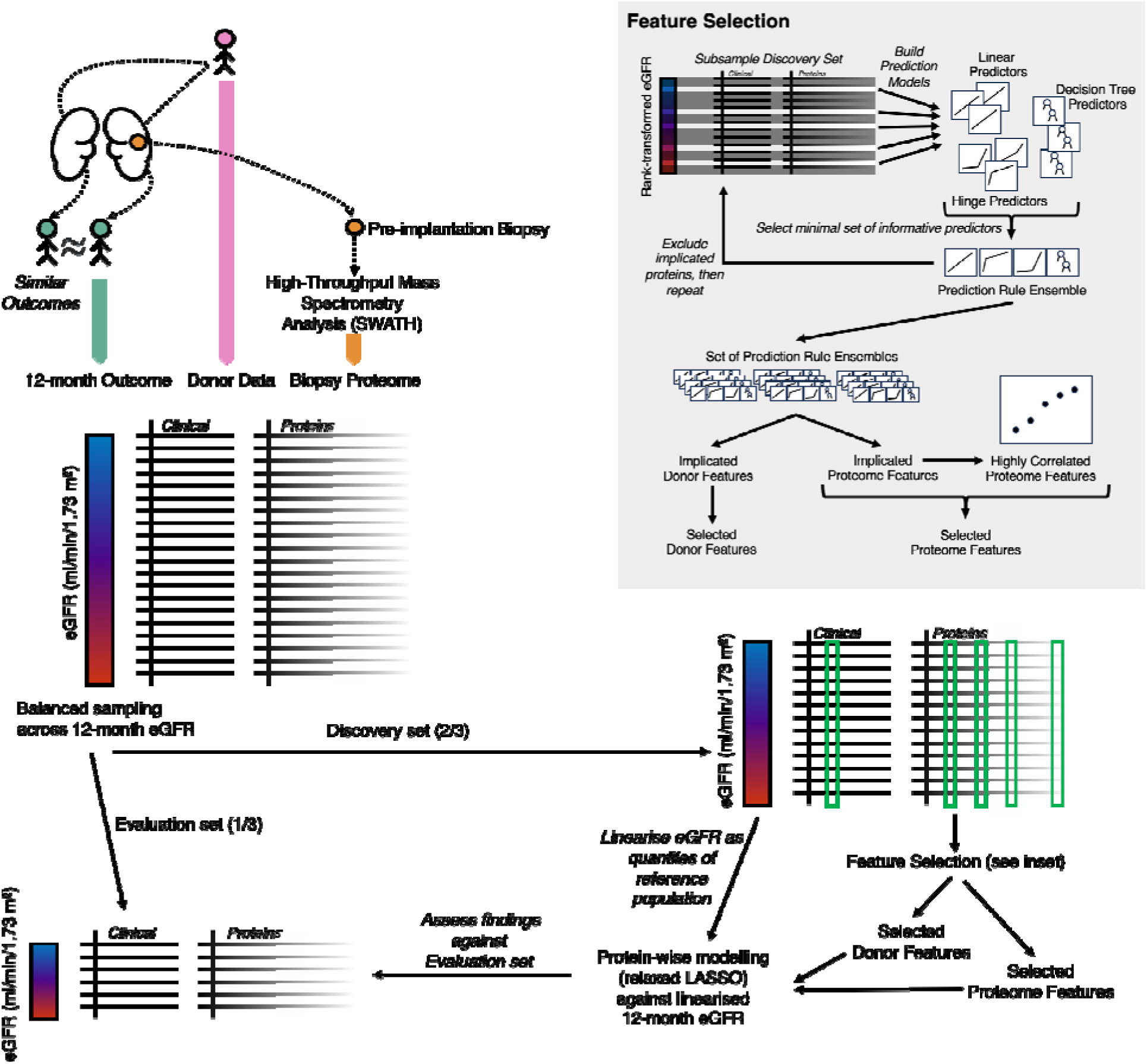
Experimental design to discover donor kidney proteome associations with transplant outcome. We selected biopsies from QUOD biobank taken from one kidney from each donor pair. Donor kidney samples were selected randomly from pairs where both recipients had similar outcomes. The biopsy samples were subjected to proteomic analysis to yield a snapshot of the organ proteome at kidney retrieval. We analyzed donor characteristics and clinical variables and protein abundances in a combined model against recipient eGFR at 12 months posttransplant (eGFR12; units given in ml/min/1.73 m^2^).

Over 2000 iterations, PRE modeling generated 3282 individual prediction rules (Fig 2A). Despite all clinical variables remaining in the dataset throughout iteration, rules were dominated by prediction functions based on donor age alone (3075/3282; 93.7%). Of the remaining rules, 194 (5.9%) referenced proteins and just over a third of this total (73) also involved donor age. These rules were predominantly decision tree and spline-based (out of 194 rules, 106 were decision trees, 20 were linear fits and 68 were spline fits), indicating that protein effects on outcome are not well-modeled by simple linear associations with protein abundance alone. There were only 13 (0.4%) rules referencing clinical variables other than donor age; of these, five referenced donor type, two referenced donor history of hypertension, one referenced hypotension preceding donor management, two referenced donor urine output in the last 24 hours, one referenced the time between circulatory arrest and cold perfusion, and two referenced recipient pretransplant dialysis length.

**Figure 2:**
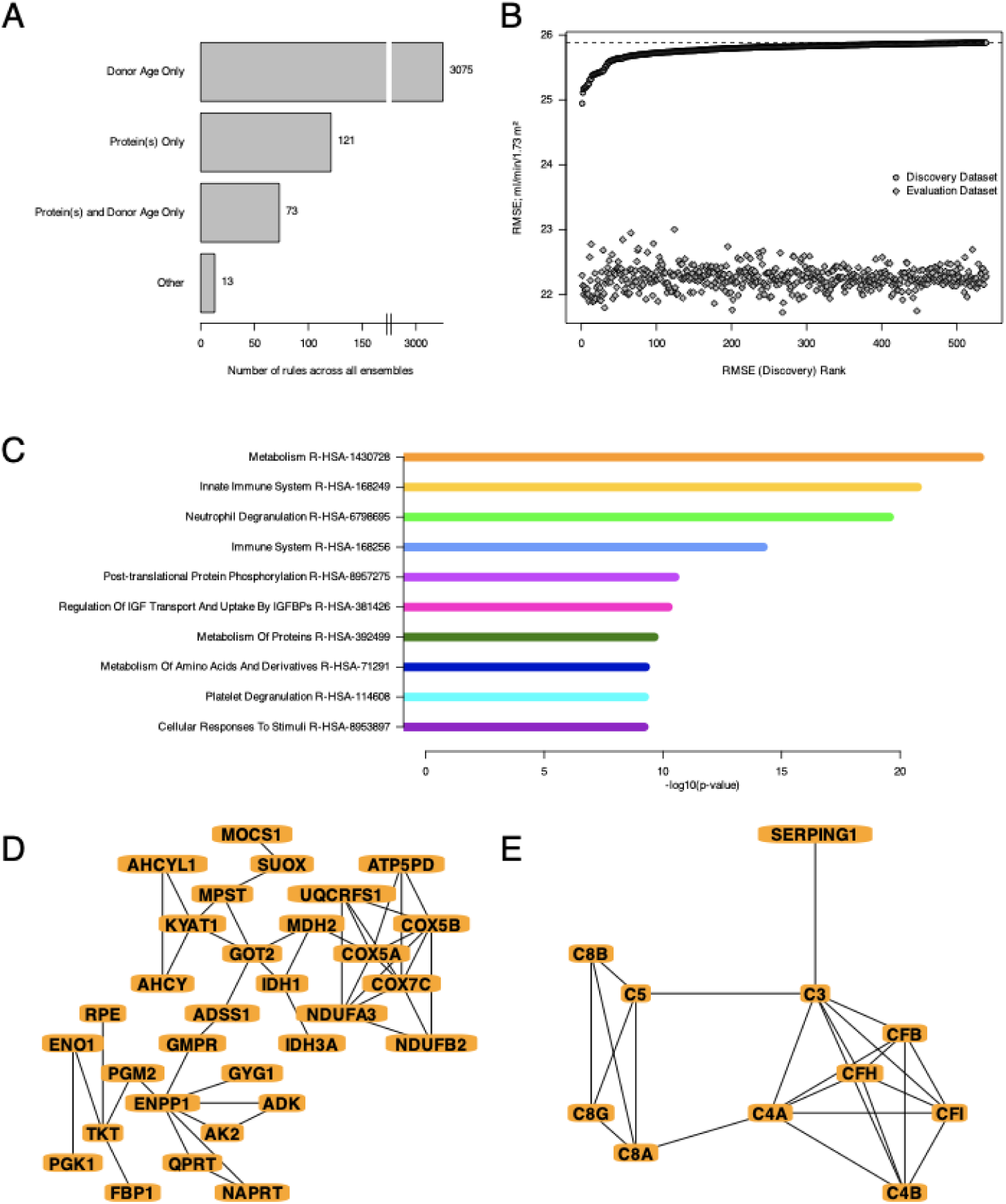
Protein association with transplant outcome is dominated by donor age effects. A: Breakdown of prediction function (‘rule’) terms identified in any ensemble across 2000 PRE iterations. B: Evaluation of individual protein model performance (Root-Mean-Squared Error, lower is better) on the discovery dataset used for fitting each model (circles) and corresponding performance on a ‘held out’ evaluation dataset (diamonds). The dashed line indicates the performance of the donor-age only model on the discovery dataset, applied as a maximum threshold for protein selection. C: Top 10 Reactome terms identified by pathway analysis of all 539 proteins. D & E: STRINGdb functional association networks of identified proteins associated with Metabolism (D) and Innate Immune System (E), the most and second most significant Reactome terms respectively. Only connected vertices are shown. Edges represent ‘highest confidence’ functional interactions (confidence score >0.9).

Based on our observations from PRE modelling, we tested individual proteins for association with eGFR12 using relaxed LASSO regression with protein, donor age, and age:protein interaction terms ^21^. We normalized eGFR12 outcomes against the distribution of eGFR12 across UK recipients between 2016-2021 (see Supplementary Methods) to linearize outcomes and avoid over-fitting to the particular outcome distribution within our discovery data. We selected only proteins which gave models better than donor age alone (see Supplementary Methods).

This approach yielded a set of 539 proteins associated with transplant outcome after accounting for the expected effect of donor age alone (donor age only; DAO model; Supplementary Table ST2), with mean root mean square error (RMSE) on our discovery dataset of 25.78 ml/min/1.73m^2^, where the upper limit (the DAO model RMSE) was 25.88 ml/min/1.73m^2^. Protein associations identified in the discovery cohort replicated well in our evaluation cohort. RMSE on this evaluation data was consistently lower (mean RMSE 22.25 ml/min/1.73m^2^; Figure 2B).

Reactome pathway analysis of the 539 outcome-associated proteins (Figure 2C) indicated that a key mechanistic theme related to metabolism (Fig 2D), including key enzymes in gluconeogenesis (Enolase 1; ENO1 and Fructose-1,6-bisphosphatase 1; FBP1) and the pentose phosphate pathway (Transketolase; TKT), members of the citric acid cycle (Glutamic-oxaloacetic transaminase 2; GOT2, Malate dehydrogenase 2; MDH2) and components of electron transport chain Complexes I (NADH:ubiquinone oxidoreductase subunits A3 and B2; NDUFA3, NDUFB2), III (Cytochrome b-c1 complex subunit 5; UQCRFS1) and IV (Cytochrome c oxidase subunits 5A, 5B and 7C; COX5A, COX5B, COX7C). A second key mechanistic theme was innate immune responses (Figure 2E), particularly regulation of complement, via both the classical pathway (Complement factor 4 and B; CF4A, CF4B) and alternative pathway (Complement factors B, H, I, 3, 5 8A, 8B and 8G; CFB, CFH, CFI, CF3, CF5, CF8A, CF8B, CF8G).

### Association of Proteins with Posttransplant Outcome is Modulated by Donor Age

For all 539 proteins, their association with outcome was also modulated by donor age (Supplementary Table ST2). To explore the implications of each protein model further, we considered how the models compared to the expected outcome using a DAO We modeled the outcomes according to donor age for each protein at low abundance (10^th^ percentile) and high abundance (90^th^ percentile) and compared the results to the DAO model, ranking proteins by log2 fold change Euclidean distance to DAO across ages (Figure 3C,D). Of the 539 proteins that outperformed the DAO model, we found that low abundance was associated with better outcome than the DAO model (Figure 3C) while high protein abundance was associated with worse outcome than the DAO model (Figure 3D). Furthermore, the amount by which high protein abundance associated with worse outcomes generally increased from age of 50 years.

**Figure 3:**
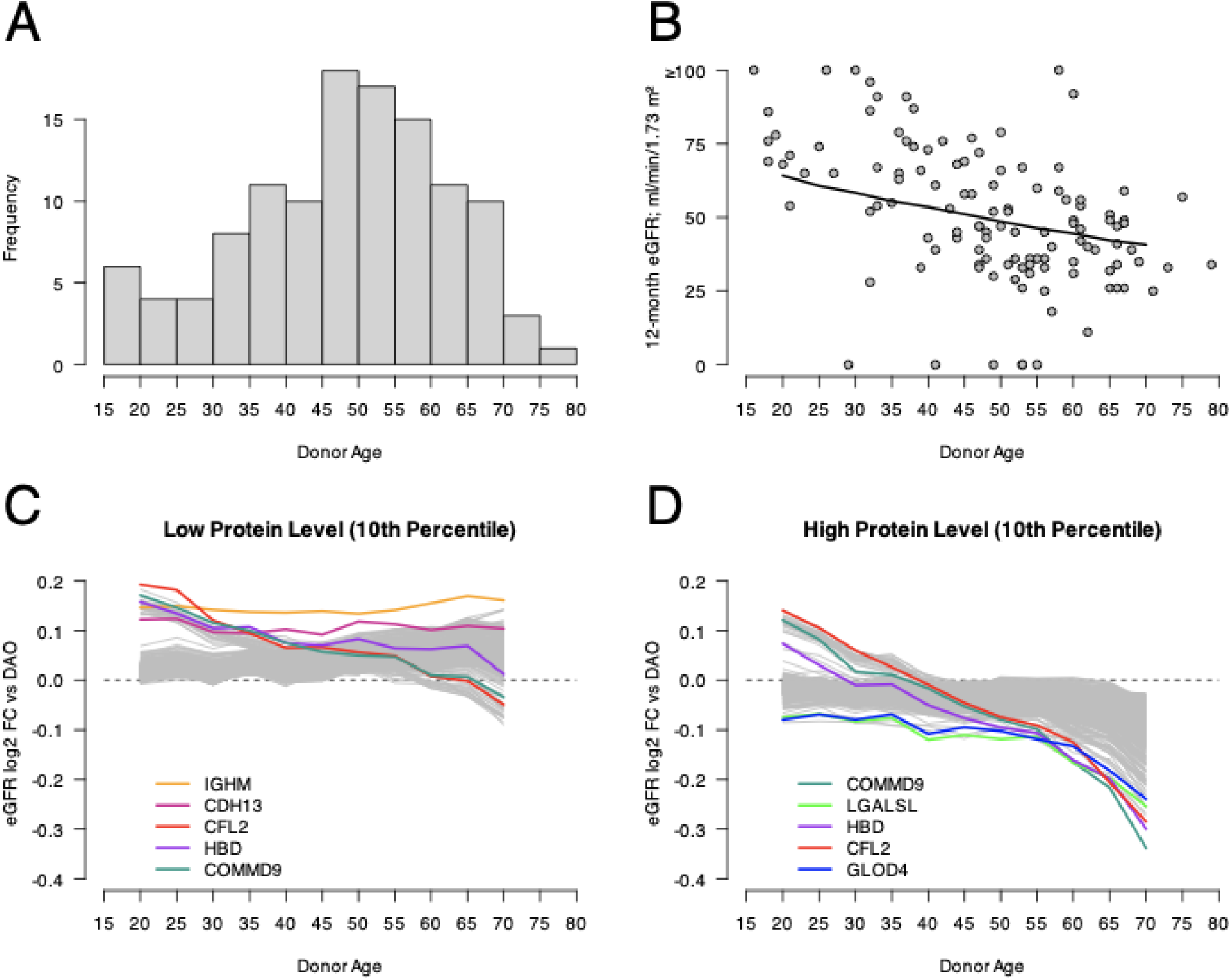
Modeled associations between proteins and kidney transplant outcome change with donor age. Our modeling found that the effect of candidate protein levels on outcome changed with age. A: Distribution of donor age over out dataset. B: Donor-age-only model; fitted effect of donor age on eGFR12. C & D: Effect of low protein abundance (10^th^ percentile) (C) and high protein abundance (90^th^ percentile) (D) on modeled eGFR12, relative to donor age-only (DAO) model eGFR12, across donor age. The 5 most different (highest Euclidean distance to donor age-only model) proteins are shown in each case.

We compared the top five proteins that diverged the most from the DAO model at low protein abundance (in order of decreasing divergence: Immunoglobulin heavy constant mu; IGHM, Cadherin 13; CDH13, Cofilin 2; CFL2, Hemoglobin subunit delta; HBD and COMM domain-containing protein 9; COMMD9) (Figure 3C) and the top five that diverged the most from the DAO model at high protein abundance (in order of decreasing divergence: COMMD9, Lectin galactoside-binding-like protein; LGALSL, HBD, CFL2 and Glyoxalase domain-containing protein 4; GLOD4) (Figure 3D), revealing three overlapping proteins (CFL2, HBD and COMMD9). These three proteins behaved similarly when expressed in donor kidney biopsies at low and high abundance, and they were associated with better outcomes than the DAO model at low age, trending towards worse outcomes as age increased (Figure 4A,B,C). At high abundance the gradient with respect to age was so steep that high abundance of these proteins associated with slightly better than otherwise expected outcome (better than the DAO model) in young donors but considerably worse than otherwise expected outcome (worse than the DAO model) in old donors.

**Figure 4:**
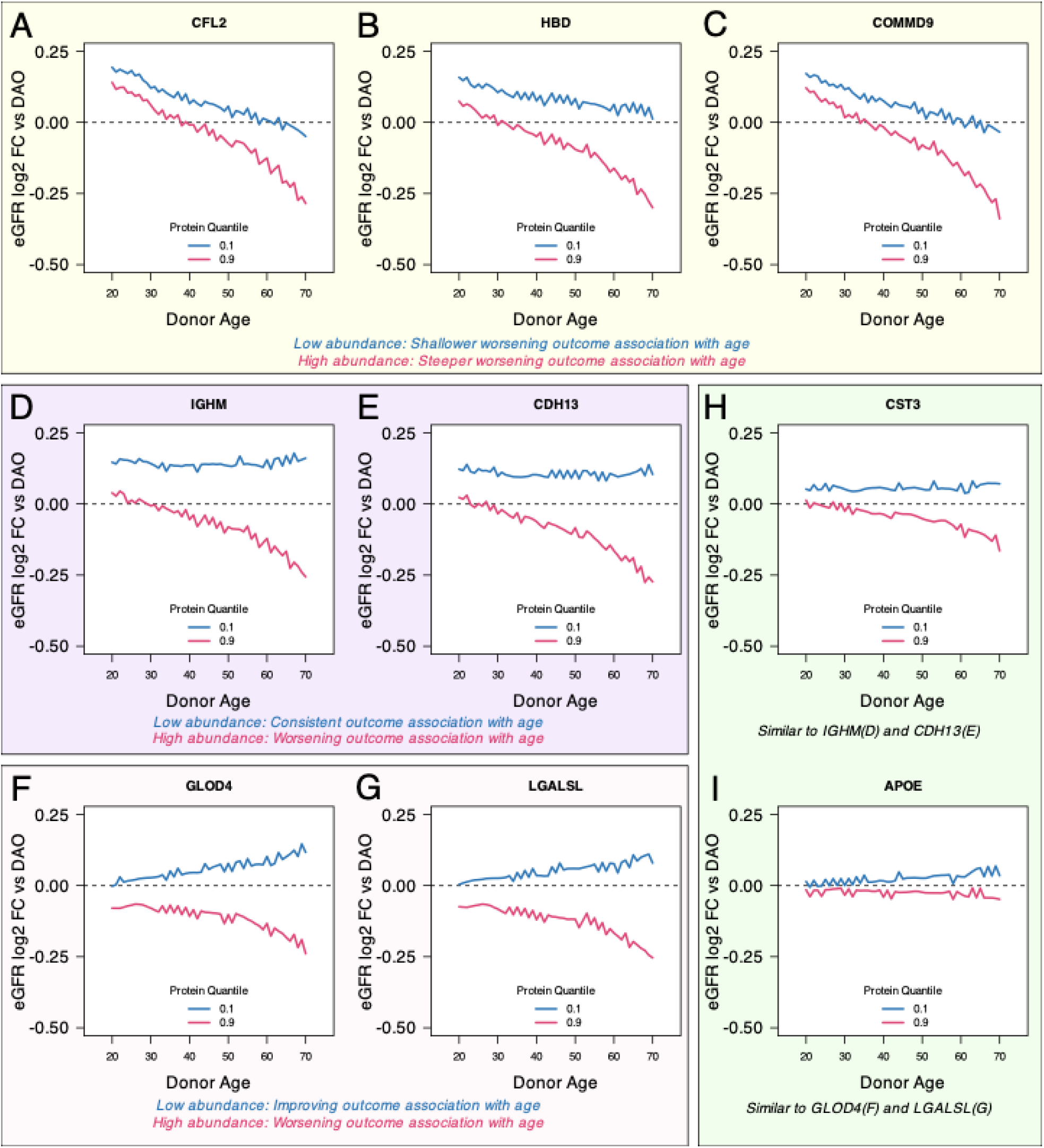
Effect of difference in protein abundances change with age. Comparisons of the effect of low and high protein abundance on modeled eGFR12, across donor age. Panels A-G (yellow, purple and pink backgrounds; qualitative grouping by relationship with age – see Results): proteins highlighted in Figure 3C,D. Panels H & I (green background): known kidney function markers. Fold changes shown (y axes) are the modeled eGFR12 relative to donor age-only (DAO) model eGFR12, across donor age. Qualitative summaries of low and high abundance effects are given below the panels.

In contrast, the other four most divergent proteins across low and high abundance (IGHM, CDH13, GLOD4, LGALSL) behaved differently in their association with outcome over age at low abundance, with protein abundance associating with better outcomes regardless of age (IGHM, CDH13; Figure 4D,E) or even slightly improving with age (GLOD4, LGALSL; Figure 4F,G).

While proteins with the most exaggerated difference from the DAO model have the strongest mechanistic implications, our list of 539 candidates also includes proteins classically associated with kidney function. Of particular note was Cystatin-C (CST3), which showed a similar relationship with age as IGHM and CDH13, i.e. relatively consistent over age at low abundance but associating with increasingly worse outcomes as age increased at high abundance (Figure 4H). Another protein associated with outcomes was Apolipoprotein E (APOE), which is also strongly associated with genetic age in other pathologies (particularly Alzheimer’s Disease). We observed modest improvement in outcomes with age at low APOE abundance, similar to GLOD4 and LGALSL (Figure 4I).

Collectively, our protein modeling findings show that, in deceased donor kidneys, even accounting for age effects on outcome, the interpretation of how proteomic changes associate with transplant outcome depends on donor age.

### Comparison to Single-Cell Transcriptome Data Suggests Protein-Outcome Associations May Localize to Specific Cell Types

To further characterize our 539 eGFR12-associated proteins, we sought to contextualize biological changes in this set of proteins as a result of organ damage. We evaluated our protein set against a publicly available scRNA-seq dataset obtained from “Normal” (living donors, healthy reference state) and “Damaged” kidneys with altered physiology due to assessed AKI or CKD ^22^. We compared the set of transcripts that matched our proteomic findings between healthy and injured kidney cell types. At 5% false discovery rate, we observed that matched transcripts showed an overall increase in cells from Damaged kidneys, across both kidney tissue and immune cell types (Figure 5, ‘All Matching Transcripts’), providing further confidence that our set of proteins with age modulated outcome associations represented omic changes caused by kidney damage. To explore these data further, we considered transcript sub-sets corresponding to the top 10 shortlisted Reactome pathways that were defined by our findings (Figure 2C). We observed that different pathways showed varying levels of transcript abundance increase across both kidney tissue and immune cell types (Figure 5). For example, proximal tubule epithelial cells showed a downregulation of transcripts associated with “*Metabolism*” and “*Metabolism Of Amino Acids And Derivatives”,* whereas other kidney tissue cell types showed upregulation of these pathways.

**Figure 5:**
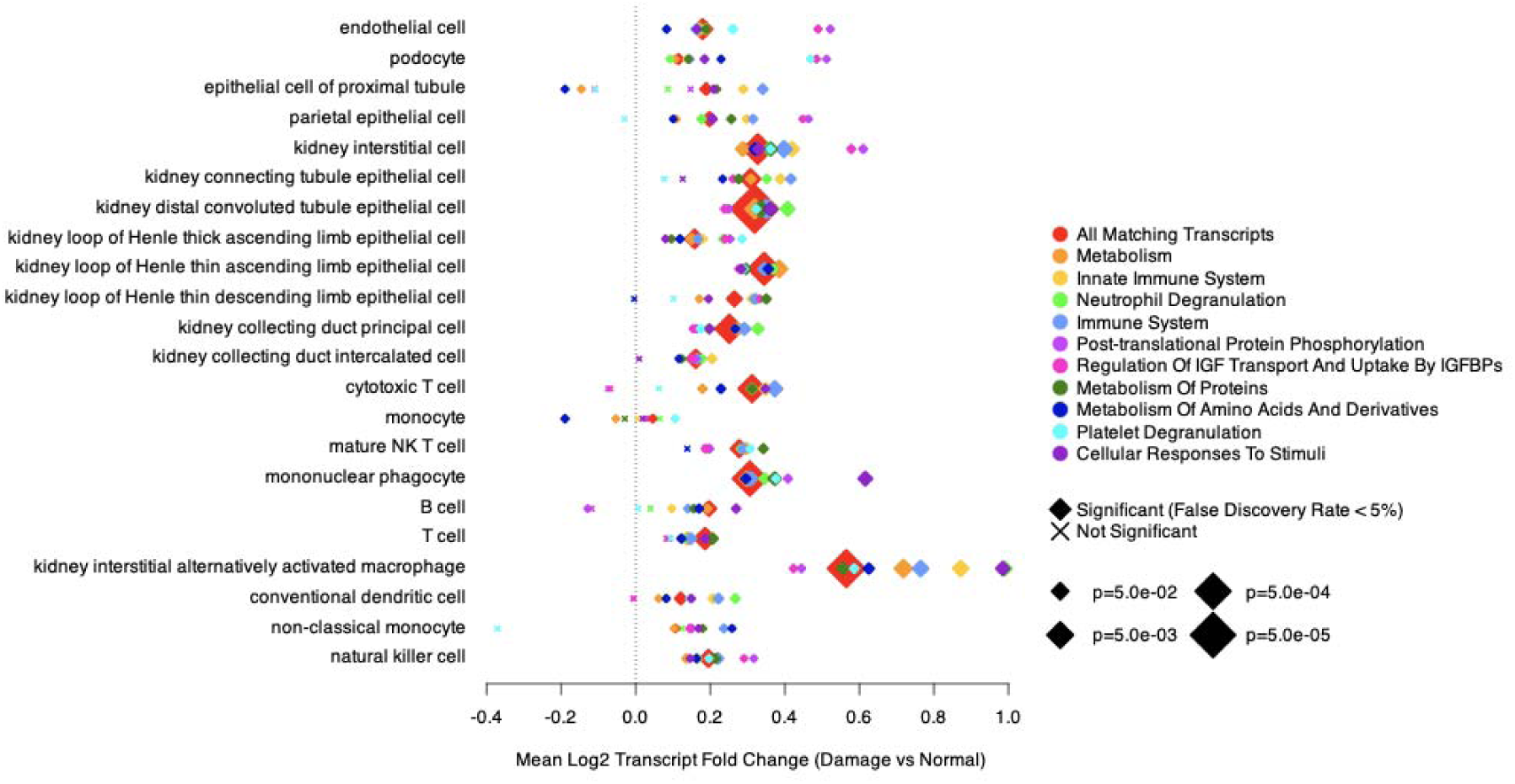
Independent scRNA-seq data confirm corresponding transcriptomic increases in injured organs. Evaluation of our protein set against independent transcriptomic data comparing Normal kidneys from living donors *versus* Damaged kidneys assessed as having AKI or CKD (Lake *et al*.) ^22^. Transcripts matching our set of 539 proteins were considered across cell type assignments in Lake *et al.* (labels on left side of panel). We considered the full set of transcripts (‘All Matching Transcripts’, red symbols), as well as subsets of protein-matching transcripts corresponding to the top 10 Reactome pathways as reported in Figure 2C. X-axis shows the mean log2 fold change across all matching transcripts within each set.

## DISCUSSION

We examined the relationship between posttransplant kidney function with pretransplant kidney proteomes by analyzing biopsies from 185 deceased donor kidneys with complete donor and recipient associated metadata. Our analyses revealed that ageing changes how proteomic profiles in donor kidneys associate with transplant outcome and that modeling kidney proteome information and donor age increases granularity in understanding donor kidney quality. Notably, we observed that, even accounting for the independent effect of donor age on outcome, interpretation of proteomic signals with respect to outcome is modified by donor age.

Within our set of 539 age modulated proteins associated with outcome there are common themes of metabolic disruption and innate immune responses, (Figure 2C,D,E). This finding is consistent with previous studies of transplant outcome ^16,17^, as well as transcriptomic analyses of diseased kidneys ^23^; our data further suggest that these associations are age-modulated. The set of proteins we identified showed increases in matching transcripts derived by comparing healthy reference (living donor) versus injured (AKI and CKD) kidneys ^22^ and differences in pathway-specific fold changes between kidney tissue cell types also suggest that specific cell types may drive particular components of our reported protein set. Future studies exploring spatial patterns of cell omic profile associations with organ susceptibility and resilience to injury insults across age have the potential to identify age-informed intervention strategies.

Our confidence in our modeling is further strengthened by detection of known kidney markers. Both Cystatin-C (CST3) and Apolipoprotein E (APOE) associate with kidney dysfunction and transplant outcomes in recipients ^24–29^. Serum CST3 is recognized as a marker of kidney function ^30,31^, including in transplantation for both recipients ^24,25,32^ and donors ^33^ since levels are linked to both glomerular filtration rate and its role as an inflammatory mediator ^34–36^. Our identification of an association between outcome and tissue CST3 further emphasizes the relevance of the inflammatory role. APOE genetic variants associate with kidney dysfunction risk ^37–39^ and have widely characterized age-dependent genetic association with other pathologies such as cardiovascular disease and Alzheimer’s Disease ^40^.

The proteins we highlight as showing the most difference from the DAO model (CFL2, COMMD9, HBD) were also consistent with the broader mechanistic theme of inflammation across the full list of proteins. Cofilins such as CFL2 have been shown to be significantly upregulated in an AKI cell model and may mediate damage via endoplasmic reticulum stress-mediated ferroptosis ^41^, linking to inflammation ^42^ and diabetic nephropathy ^43^. COMMD9 associates with inflammatory response via the NF-kB pathway ^44^. HBD is a relatively small component of adult hemoglobin which shows increased expression in inflammatory conditions ^45,46^; previous studies have identified HBD as a marker for early diabetic kidney disease ^47^. Since HBD is a blood component, further investigation is required to determine whether the signal is due to inadequate kidney flushing variance that occurs prior to the biopsy being taken, and if so whether this is purely a protocol artefact or a true signal, possibly driven by kidney microvasculature effects that impair flushing.

Donor complement activation as part of the innate immune response and inflammation is associated with both acute rejection ^48–50^ and inferior 12-month outcome^51^. Existing evidence and our findings particularly indicate inflammation mediated by the anaphylatoxins C3A, C4B, C5A ^52^, suggesting an increase in immunogenicity with subsequent negative impact on transplant outcomes. Preventing inflammation and associated damage by anti-complement interventions in the donor, or during organ preservation (as demonstrated successfully in mouse models ^53^) may therefore be a route to improve outcomes ^54,55^.

Our other highlighted proteins are also have been linked with patterns of kidney injury. GLOD4 regulates methylgloxal, accumulation of which drives renal injury in diabetic-drive hyperglycemia ^56^, LGALSL encodes a paralog of Galectin 9, associated with autoimmune renal damage^57^, and differences in IGHM and CDH13 expression have previously been linked to renal damage in mouse models (acute-chronic injury transition ^58^ and ischemia-reperfusion injury ^59^ respectively).

Organ allocation algorithms impose a close link between donor and recipient age in the sample cohort, so while we might interpret these age-moderated effects as an increase in organ susceptibility with donor age, it could also represent a greater ability to repair a given level of damage in younger recipients, although the observation that transplant of older organs induces ageing-like senescence in young recipients in a mouse model ^60^ implies that donor age is highly relevant to transplant outcome even in young recipients. Additionally, we consider only chronological donor age, rather than a representation of the epigenomic biological clock ^61^, or organ-specific ageing ^62,63^, both of which may account for some variation observed with respect to both donors and recipients. In contrast to donor age, a clinical variable notable by its absence from our association findings was donor type ^64^, which appeared in just 5 (0.2%) of the initial feature selection rules. Previous transcriptomic comparisons of DBD and DCD biopsies to living donors found very similar expression profiles across donor type except in samples taken after reperfusion ^50^, suggesting that donor type specific biological stress may manifest post reperfusion.

In our multicentered collected cohort, our protein models generalized well to samples unseen during model training (Figure 2B), reproduced known markers of kidney damage among many new leads and were consistent with independent kidney damage transcriptome data. The observed effect of donor age on modulating kidney proteomes contributes to our understanding of molecular mechanisms of kidney injury and offers new approaches in the development of improved assessment tools to stratify transplant risk from older donors. In particular, our findings suggest that “one size fits all” interventions will continue to be ineffective for many cases, until molecular signals interpreted in the light of donor age are given wider consideration to guide personalized decision-making.

A follow up larger study would have increased power to deconvolute age and protein effects, especially if paired with variant sequencing to understand genetic diversity, and explore the potential developing protein biomarkers for donor kidney assessment.

## Data Availability

The mass spectrometry proteomics data have been deposited to the ProteomeXchange Consortium via the PRIDE 67 partner repository with the dataset identifier PXD033428. All other data produced are contained in the manuscript or supplemental materials.

https://www.ebi.ac.uk/pride/

## SUPPLEMENTARY MATERIALS

Supplementary Methods: Technical protocols for sample extraction, mass spectrometry deep proteome analysis of samples and statistical analysis.

Supplementary Table ST1: Clinical variable p-values for association with donor type and outcome, and summary of clinical value imputation.

Supplementary Table ST2: Summary of results for all proteins identified as associating with eGFR12.

Note that coefficient values provided are for linearized eGFR12, i.e. they relate to eGFR12 quantiles ranging from 0 to 1.

Supplementary Figure SF1: Summary of protein quantitation quality. A: Percentage of missing values across all proteins, showing the cut-off (red line) for poorly quantified proteins excluded from analysis. B: Correlation between quantitation values for each of the 3 sets of paired kidneys.

Supplementary Figure SF2: Summary of protein quantitation imputation, showing the fraction of imputed values across distribution of all values.

## DATA AND MATERIALS AVAILABILITY

The mass spectrometry proteomics data have been deposited to the ProteomeXchange Consortium via the PRIDE ^65^ partner repository with the dataset identifier PXD033428.

## AUTHORSHIP PAGE

### Author contributions

Conceptualization: MK

Methodology: PDC, RV, SF, SD, RF, BMK, AS, ES, RJP, MK

Investigation: PDC, RV, SF, PJ, SD, IV, KT, AS

Visualization: PDC

Funding acquisition: BMK, RJP, MK

Project oversight: MK

Supervision: RF, AS, MK

Writing – original draft: PDC, ES, MK

Writing – review & editing: All authors

### Disclosure

No conflicts of interest to disclose by any of the authors.

### Funding

This study was supported by NHS Blood and Transplant funding awarded to MK. SF was supported by Kidney Research UK, grant reference KS_RP_002_20210111 awarded to MK. PDC was supported by Chinese Academy of Medical Sciences 2018-I2M-2-002 awarded to BMK.

## ABBREVIATIONS

AKI: Acute Kidney Injury
CKD: Chronic Kidney Disease
DAO: Donor Age-Only
DBD: Donation after Brain Death
DCD: Donation after Circulatory Death
eGFR: Estimated Glomerular Filtration Rate
eGFR12: Estimated Glomerular Filtration Rate of recipient at 12 months posttransplant
ESKD: End-Stage Kidney Disease
LASSO: Least Absolute Shrinkage and Selection Operator; a regularized regression technique
PRE: Prediction Rule Ensemble
RMSE: Root Mean Squared Error
scRNA-seq: Single-cell Transcriptomics
QUOD: Quality in Organ Donation (biobank)

## ACKNOWLEDGMENTS

We thank the UK QUOD Consortium and NHS Blood and Transplant UK Registry for providing the samples and the associated clinical and demographic metadata.; in particular we thank Sheba Ziyenge, Lewis Simmonds, Dr Meng Sun and Dr Sarah Cross, Dr Sergei Maslau and Mr Tomas Surik for their support on the QUOD sample selection.

We thank members of the Discovery Proteomics Facility within the TDI Mass Spectrometry Laboratory for expert help with mass spectrometry analysis, and members of the Lindgren and Nellåker groups at the BDI for informative discussions regarding statistical modeling.

## Notes

### Competing Interest Statement

The authors have declared no competing interest.

### Funding Statement

This study was funded by NHS Blood and Transplant WP15-07 awarded to MK & RJP. SF was supported by Kidney Research UK, grant reference KS_RP_002_20210111 awarded to MK. PDC was supported by a Chinese Academy of Medical Sciences 2018-I2M-2-002 awarded to BMK.

### Author Declarations

NHS Health Authority, National Research Ethics Service, gave ethical approval for IRAS project ID: 87824, Quality in Organ Donation (QUOD) (NW/18/0187), sponsored by University of Oxford

### Summary of Updates

Revision to the analysis and presentation of Figure 5, to improve result granularity. Transcriptomics analysis now relies on the CELLxGENE Seurat-encapsulated count data rather than the relative abundance data obtained via the web portal, since that is no longer easily retrieved in bulk.

## REFERENCES

1. Callaghan CJ, Mumford L, Pankhurst L, Baker RJ, Bradley JA, Watson CJE. Early Outcomes of the New UK Deceased Donor Kidney Fast-Track Offering Scheme. Transplantation. 2017;101(12):2888–2897. doi:10.1097/TP.0000000000001860

2. Summers DM, Johnson RJ, Hudson AJ, et al. Standardized deceased donor kidney donation rates in the UK reveal marked regional variation and highlight the potential for increasing kidney donation: a prospective cohort study†. Br J Anaesth. 2014;113(1):83–90. doi:10.1093/bja/aet473

3. Denic A, Glassock RJ, Rule AD. The Kidney in Normal Aging: A Comparison with Chronic Kidney Disease. Clin J Am Soc Nephrol. 2022;17(1):137–139. doi:10.2215/CJN.10580821

4. O’Hare AM, Choi AI, Bertenthal D, et al. Age affects outcomes in chronic kidney disease. J Am Soc Nephrol. 2007;18(10):2758–2765. doi:10.1681/ASN.2007040422

5. Ishani A, Xue JL, Himmelfarb J, et al. Acute Kidney Injury Increases Risk of ESRD among Elderly. J Am Soc Nephrol. 2009;20(1):223–228. doi:10.1681/ASN.2007080837

6. Ibrahim M, Greenhall GHB, Summers DM, et al. Utilization and outcomes of single and dual kidney transplants from older deceased donors in the United Kingdom. Clin J Am Soc Nephrol. 2020;15(9):1320–1329. doi:10.2215/CJN.02060220

7. Rao PS, Schaubel DE, Guidinger MK, et al. A comprehensive risk quantification score for deceased donor kidneys: the kidney donor risk index. Transplantation. 2009;88(2):231–236. doi:10.1097/TP.0b013e3181ac620b

8. Watson CJE, Johnson RJ, Birch R, Collett D, Bradley JA. A Simplified Donor Risk Index for Predicting Outcome After Deceased Donor Kidney Transplantation. Transplantation. 2012;93(3):314–318. doi:10.1097/TP.0b013e31823f14d4

9. Neuberger J, Callaghan C. Organ utilization – the next hurdle in transplantation? Transpl Int. 2020;33(12):1597–1609. doi:10.1111/tri.13744

10. Clayton PA, Dansie K, Sypek MP, et al. External validation of the US and UK kidney donor risk indices for deceased donor kidney transplant survival in the Australian and New Zealand population. Nephrol Dial Transplant. 2019;34(12):2127–2131. doi:10.1093/NDT/GFZ090

11. Aubert O, Reese PP, Audry B, et al. Disparities in acceptance of deceased donor kidneys between the United States and France and estimated effects of increased US acceptance. JAMA Intern Med. 2019;179(10):1365–1374. doi:10.1001/jamainternmed.2019.2322

12. Yu K, Husain SA, King K, Stevens JS, Parikh CR, Mohan S. Kidney nonprocurement in deceased donors with acute kidney injury. Clin Transplant. Published online August 4, 2022:e14788. doi:10.1111/ctr.14788

13. Hall IE, Akalin E, Bromberg JS, et al. Deceased-donor acute kidney injury is not associated with kidney allograft failure. Kidney Int. 2019;95(1):199–209. doi:10.1016/j.kint.2018.08.047

14. Liu C, Hall IE, Mansour S, Thiessen Philbrook HR, Jia Y, Parikh CR. Association of Deceased Donor Acute Kidney Injury With Recipient Graft Survival. JAMA Network Open. 2020;3(1):e1918634. doi:10.1001/jamanetworkopen.2019.18634

15. Mansour SG, Khoury N, Kodali R, et al. Clinically adjudicated deceased donor acute kidney injury and graft outcomes. PLoS One. 2022;17(3):e0264329. doi:10.1371/journal.pone.0264329

16. Kaisar M, Van Dullemen L, Charles P, et al. Subclinical Changes in Deceased Donor Kidney Proteomes Are Associated with 12-month Allograft Function Posttransplantation-A Preliminary Study. Transplantation. 2019;103(2):323–328. doi:10.1097/TP.0000000000002358

17. Zhang R, Trotter PB, McCaffrey J, et al. Assessment of biological organ age using molecular pathology in pre-transplant kidney biopsies. Kidney Int. Published online April 29, 2024. doi:10.1016/j.kint.2024.03.028

18. von Moos S, Akalin E, Mas V, Mueller TF. Assessment of Organ Quality in Kidney Transplantation by Molecular Analysis and Why It May Not Have Been Achieved, Yet. Front Immunol. 2020;11:833. doi:10.3389/fimmu.2020.00833

19. Raynaud M, Aubert O, Divard G, et al. Dynamic prediction of renal survival among deeply phenotyped kidney transplant recipients using artificial intelligence: an observational, international, multicohort study. The Lancet Digital Health. 2021;3(12):e795–e805. doi:10.1016/S2589-7500(21)00209-0

20. Fokkema M. Fitting prediction rule ensembles with R package pre. J Stat Softw. 2020;92(1):1–30. doi:10.18637/jss.v092.i12

21. Friedman JH, Hastie T, Tibshirani R. Regularization Paths for Generalized Linear Models via Coordinate Descent. J Stat Softw. 2010;33:1–22. doi:10.18637/jss.v033.i01

22. Lake BB, Menon R, Winfree S, et al. An atlas of healthy and injured cell states and niches in the human kidney. Nature. 2023;619(7970):585–594. doi:10.1038/s41586-023-05769-3

23. Abedini A, Levinsohn J, Klötzer KA, et al. Single-cell multi-omic and spatial profiling of human kidneys implicates the fibrotic microenvironment in kidney disease progression. Nat Genet. Published online July 24, 2024:1–13. doi:10.1038/s41588-024-01802-x

24. Coll E, Botey A, Alvarez L, et al. Serum cystatin C as a new marker for noninvasive estimation of glomerular filtration rate and as a marker for early renal impairment. Am J Kidney Dis. 2000;36(1):29–34. doi:10.1053/ajkd.2000.8237

25. Christensson A, Ekberg J, Grubb A, Ekberg H, Lindström V, Lilja H. Serum cystatin C is a more sensitive and more accurate marker of glomerular filtration rate than enzymatic measurements of creatinine in renal transplantation. Nephron Physiol. 2003;94(2):19–27. doi:10.1159/000071287

26. Rodrigo E, Ruiz JC, Fernández-Fresnedo G, et al. Cystatin C and albuminuria as predictors of long-term allograft outcomes in kidney transplant recipients. Clin Transplant. 2013;27(2):E177–83. doi:10.1111/ctr.12082

27. Kahraman S, Kiykim AA, Altun B, et al. Apolipoprotein E gene polymorphism in renal transplant recipients: effects on lipid metabolism, atherosclerosis and allograft function. Clin Transplant. 2004;18(3):288–294. doi:10.1111/j.1399-0012.2004.00162.x

28. Hernández D, Salido E, Linares J, et al. Role of apolipoprotein E epsilon 4 allele on chronic allograft nephropathy after renal transplantation. Transplant Proc. 2004;36(10):2982–2984. doi:10.1016/j.transproceed.2004.10.038

29. Cofán F, Cofan M, Rosich E, et al. Effect of apolipoprotein E polymorphism on renal transplantation. Transplant Proc. 2007;39(7):2217–2218. doi:10.1016/j.transproceed.2007.06.011

30. Kyhse-Andersen J, Schmidt C, Nordin G, et al. Serum cystatin C, determined by a rapid, automated particle-enhanced turbidimetric method, is a better marker than serum creatinine for glomerular filtration rate. Clin Chem. 1994;40(10):1921–1926. https://www.ncbi.nlm.nih.gov/pubmed/7923773

31. Newman DJ, Thakkar H, Edwards RG, et al. Serum cystatin C measured by automated immunoassay: a more sensitive marker of changes in GFR than serum creatinine. Kidney Int. 1995;47(1):312–318. doi:10.1038/ki.1995.40

32. Lima JR, Salgado JV, Ferreira TC, Oliveira MI, dos Santos AM, Filho NS. Cystatin C and inflammatory markers in kidney transplant recipients. Rev Assoc Médica Bras (Engl Ed). 2011;57(3):341–346. doi:10.1016/s2255-4823(11)70070-4

33. Michelakis I, Fawaz S, Vaughan R, et al. #1645 The role of donor Cystatin-C in posttransplant outcomes in kidney transplantation. Nephrol Dial Transplant. 2024;39(Supplement_1):gfae069–0175-1645. doi:10.1093/ndt/gfae069.175

34. Singh D, Whooley MA, Ix JH, Ali S, Shlipak MG. Association of cystatin C and estimated GFR with inflammatory biomarkers: the Heart and Soul Study. Nephrol Dial Transplant. 2007;22(4):1087–1092. doi:10.1093/ndt/gfl744

35. Okura T, Jotoku M, Irita J, et al. Association between cystatin C and inflammation in patients with essential hypertension. Clin Exp Nephrol. 2010;14(6):584–588. doi:10.1007/s10157-010-0334-8

36. Potok OA, Ix JH, Shlipak MG, et al. Cystatin C– and Creatinine-Based Glomerular Filtration Rate Estimation Differences and Muscle Quantity and Functional Status in Older Adults: The Health, Aging, and Body Composition Study. Kidney Med. 2022;4(3):100416. doi:10.1016/j.xkme.2022.100416

37. Czaplińska M, Ćwiklińska A, Sakowicz-Burkiewicz M, et al. Apolipoprotein e gene polymorphism and renal function are associated with apolipoprotein e concentration in patients with chronic kidney disease. Lipids Health Dis. 2019;18(1):1–9. doi:10.1186/s12944-019-1003-x

38. Hsu CC, Kao WHL, Coresh J, et al. Apolipoprotein E and progression of chronic kidney disease. J Am Med Assoc. 2005;293(23):2892–2899. doi:10.1001/jama.293.23.2892

39. Saito T, Ishigaki Y, Oikawa S, Yamamoto TT. Etiological significance of apolipoprotein E mutations in lipoprotein glomerulopathy. Trends Cardiovasc Med. 2002;12(2):67–70. doi:10.1016/S1050-1738(01)00148-7

40. Liu CC, Kanekiyo T, Xu H, Bu G. Apolipoprotein E and Alzheimer disease: risk, mechanisms, and therapy. Nat Rev Neurol. 2013;9(2):106–118. doi:10.1038/nrneurol.2012.263

41. Lin S, Wang J, Cao B, Huang Y, Sheng X, Zhu Y. Cofilin-1 induces acute kidney injury via the promotion of endoplasmic reticulum stress-mediated ferroptosis. Hum Cell. 2023;36(6):1928–1937. doi:10.1007/s13577-023-00949-9

42. Chen Y, Fang ZM, Yi X, Wei X, Jiang DS. The interaction between ferroptosis and inflammatory signaling pathways. Cell Death Dis. 2023;14(3):205. doi:10.1038/s41419-023-05716-0

43. Li L, Dai Y, Ke D, et al. Ferroptosis: new insight into the mechanisms of diabetic nephropathy and retinopathy. Front Endocrinol. 2023;14:1215292. doi:10.3389/fendo.2023.1215292

44. Laulumaa S, Varjosalo M. Commander Complex-A Multifaceted Operator in Intracellular Signaling and Cargo. Cells. 2021;10(12). doi:10.3390/cells10123447

45. Kobayashi S, Ito A, Okuzaki D, et al. Expression profiling of PBMC-based diagnostic gene markers isolated from vasculitis patients. DNA Res. 2008;15(4):253–265. doi:10.1093/dnares/dsn014

46. Brunyanszki A, Erdelyi K, Szczesny B, et al. Upregulation and Mitochondrial Sequestration of Hemoglobin Occur in Circulating Leukocytes during Critical Illness, Conferring a Cytoprotective Phenotype. Mol Med. 2015;21(1):666–675. doi:10.2119/molmed.2015.00187

47. Golea-Secara A, Munteanu C, Sarbu M, et al. Urinary proteins detected using modern proteomics intervene in early type 2 diabetic kidney disease – a pilot study. Biomark Med. 2020;14(16):1521–1536. doi:10.2217/bmm-2020-0308

48. Damman J, Seelen MA, Moers C, et al. Systemic complement activation in deceased donors is associated with acute rejection after renal transplantation in the recipient. Transplantation. 2011;92(2):163–169. doi:10.1097/TP.0b013e318222c9a0

49. Damman J, Daha MR, van Son WJ, Leuvenink HG, Ploeg RJ, Seelen MA. Crosstalk between complement and Toll-like receptor activation in relation to donor brain death and renal ischemia-reperfusion injury: Complement and TLRs in kidney transplantation. Am J Transplant. 2011;11(4):660–669. doi:10.1111/j.1600-6143.2011.03475.x

50. Damman J, Bloks VW, Daha MR, et al. Hypoxia and Complement-and-Coagulation Pathways in the Deceased Organ Donor as the Major Target for Intervention to Improve Renal Allograft Outcome. Transplantation. 2015;99(6):1293–1300. doi:10.1097/TP.0000000000000500

51. van Leeuwen LL, Spraakman NA, Brat A, et al. Proteomic analysis of machine perfusion solution from brain dead donor kidneys reveals that elevated complement, cytoskeleton and lipid metabolism proteins are associated with 1-year outcome. Transpl Int. 2021;34(9):1618–1629. doi:10.1111/tri.13984

52. Dunkelberger JR, Song WC. Complement and its role in innate and adaptive immune responses. Cell Res. 2010;20(1):34–50. doi:10.1038/cr.2009.139

53. Lewis AG, Köhl G, Ma Q, Devarajan P, Köhl J. Pharmacological targeting of C5a receptors during organ preservation improves kidney graft survival: C5aR targeting improves kidney graft survival. Clin Exp Immunol. 2008;153(1):117–126. doi:10.1111/j.1365-2249.2008.03678.x

54. Biglarnia AR, Huber-Lang M, Mohlin C, Ekdahl KN, Nilsson B. The multifaceted role of complement in kidney transplantation. Nat Rev Nephrol. 2018;14(12):767–781. doi:10.1038/s41581-018-0071-x

55. van Zanden JE, Jager NM, Daha MR, Erasmus ME, Leuvenink HGD, Seelen MA. Complement therapeutics in the multi-organ donor: Do or don’t? Front Immunol. 2019;10:329. doi:10.3389/fimmu.2019.00329

56. Schumacher D, Morgenstern J, Oguchi Y, et al. Compensatory mechanisms for methylglyoxal detoxification in experimental & clinical diabetes. Mol Metab. 2018;18:143–152. doi:10.1016/j.molmet.2018.09.005

57. Zeggar S, Watanabe KS, Teshigawara S, et al. Role of Lgals9 Deficiency in Attenuating Nephritis and Arthritis in BALB/c Mice in a Pristane-Induced Lupus Model. Arthritis Rheumatol. 2018;70(7):1089–1101. doi:10.1002/art.40467

58. Lin YH, Platt MP, Fu H, et al. Global Proteome and Phosphoproteome Characterization of Sepsis-induced Kidney Injury. Mol Cell Proteomics. 2020;19(12):2030–2047. doi:10.1074/mcp.RA120.002235

59. Tsugawa-Shimizu Y, Fujishima Y, Kita S, et al. Increased vascular permeability and severe renal tubular damage after ischemia-reperfusion injury in mice lacking adiponectin or T-cadherin. Am J Physiol Endocrinol Metab. 2021;320(2):E179–E190. doi:10.1152/ajpendo.00393.2020

60. Iske J, Roesel MJ, Martin F, et al. Transplanting old organs promotes senescence in young recipients. Am J Transplant. 2024;24(3):391–405. doi:10.1016/j.ajt.2023.10.013

61. Horvath S, Raj K. DNA methylation-based biomarkers and the epigenetic clock theory of ageing. Nat Rev Genet. 2018;19(6):371–384. doi:10.1038/s41576-018-0004-3

62. Nie C, Li Y, Li R, et al. Distinct biological ages of organs and systems identified from a multi-omics study. Cell Rep. 2022;38(10):110459. doi:10.1016/j.celrep.2022.110459

63. Shen X, Wang C, Zhou X, et al. Nonlinear dynamics of multi-omics profiles during human aging. Nat Aging. Published online August 14, 2024. doi:10.1038/s43587-024-00692-2

64. Gill J, Rose C, Lesage J, Joffres Y, Gill J, O’Connor K. Use and Outcomes of Kidneys from Donation after Circulatory Death Donors in the United States. J Am Soc Nephrol. 2017;28(12):3647–3657. doi:10.1681/asn.2017030238

65. Perez-Riverol Y, Csordas A, Bai J, et al. The PRIDE database and related tools and resources in 2019: improving support for quantification data. Nucleic Acids Res. 2019;47(D1):D442–D450. doi:10.1093/nar/gky1106

