## Supplementary Methods and Figures for "Age modulates protein-outcome associations in deceased donor kidneys"

**Protein Extraction from Renal Tissue Specimens**

Deceased donor biopsies were procured, handled and stored according to consistent, predefined collection protocols designed to minimize pre-analytical variability. Donor kidney biopsies were collected ex situ immediately after flush-out and procurement of the kidney in the donor hospital on the back-table. Biopsies were obtained from the upper pole of the donor kidney cortex during back table preparation, using a 23mm needle biopsy gun. The obtained biopsies were divided in two, with one half stored in RNAlater (Thermo Scientific, Illinois, USA), then liquid nitrogen and the other half stored in formalin.

RNAlater biopsy samples were homogenized in RIPA buffer (89900, Thermo Scientific, Illinois, USA) with protease and phosphatase inhibitor cocktail (1861280, Thermo Scientific, Illinois, USA) using a bead beater (Biorad, Hertfordshire, UK) at 6500rpm for three cycles of 40 seconds with intermediate cooling on wet ice between cycles. Biopsy protein concentration was determined using a Pierce Bicinchoninic Acid protein assay kit (23227 Thermo Scientific, Illinois, USA).

**In-Solution Trypsin Digestion**

50 µg of protein homogenates were prepared. Disulfide bonds were reduced by adding 200 mM of DTT (Sigma) to a final concentration of 5 mM for 60 mins at room temperature. Free cysteine residues were alkylated by adding 200 mM of iodoacetamide (Sigma) to a final concentration of 20 mM and incubated for 60 mins at RT in the dark.

The samples were topped up to 200 µl with 6 M urea, 100 mM TrisHCl pH 8.5. Methanol/chloroform protein precipitation was used to remove detergents before tryptic digestion. In brief, 600 µl of Methanol and 150 µl of Chloroform were added and mixed. Then 450 µl of MilliQ-H2O was added and then centrifuged for 1min at 12,000 g. The upper aqueous layer was carefully removed without disturbing formed protein pellets between layers and 450 µl of Methanol was added and then centrifuged at 12,000 g for 5 min. The supernatant was removed and the protein pellets resuspended in 50 µl of 6 M Urea, 100 mM TrisHCl, pH 8.5. Urea concentration was reduced to 1 M by adding 250 µl of MillQ-H2O. Samples were digested at 37 °C overnight with Trypsin (Promega, UK) added at a 1:50 ratio (trypsin:protein). Tryptic peptides were acidified and purified using Sep-Pak C18 cartridges (WAT020515, Waters, Wilmslow, UK) and dried by Speed Vac centrifugation. Pellets were resuspended in 80 µl of resuspension buffer A (98 % MilliQ-H2O, 2 % acetonitrile 0.1 % formic acid) for LC-MS/MS analysis.

**Mass Spectrometry Analysis**

*Generation of Fractionated Pool for Spectral Library Reference*

A highly fractionated spectral library was generated as a standard reference for the subsequent analysis of individual samples. To create this spectral library, a pool sample was prepared by combining 2 µl of each tryptic digest sample prepared above. Fractionation of the pooled sample was performed using offline high-pH reverse-phase HPLC on an XBridge BEH C18 XP column (3 × 150 mm, 2.5 μm pore size, Waters no. 186006710) over a 100-minute gradient (Buffer A: water, pH10 with ammonium hydroxide. Buffer B: 90 % acetonitrile, 10 % water, pH10 with ammonium hydroxide) with fractions collected every 2 minutes. The fractions of the pool sample for the spectral library were analyzed by nanoLC-MS/MS using a Dionex Ultimate 3000 using a 75 µm x 500 mm (2 µm particle size) C18 EASY-Spray column at 250 nL/min (Thermo Scientific), coupled to an Orbitrap Fusion Lumos mass spectrometer. Peptides were separated using a 60-minute linear gradient from 2-35 % buffer B (Buffer A: 5 % DMSO, 0.1 % formic acid, 94.9% water. Buffer B: 5 % DMSO, 0.1 % formic acid, 94.9% acetonitrile). The samples were analyzed on the mass spectrometer in Data-Dependent Acquisition mode. MS1 scans were acquired in the Orbitrap with an m/z range of 400 – 1500 m/z at a resolution of 120,000 and an AGC target of 4 x 10^5^. Precursors between charge states 2+ and 7+ were selected for HCD fragmentation using the Advanced Precursor Determination option with an intensity threshold of 2.5 x 10^4^. Selected precursors were isolated using the quadrupole with a 1.6 m/z isolation window and fragmented using HCD with a normalized collision energy of 30%. MS2 spectra were acquired in the Orbitrap using a resolution of 30,000, a maximum injection time of 54 ms and an AGC target of 5 x 10^4^.

*LC-MS/MS Analysis of Individual Biological Samples*

The individual samples were analyzed on the same nanoLC-MS/MS system as above. A 45-minute linear gradient was used from 2-35% buffer B with the same buffer composition as above. In contrast to the pool samples, the individual samples were analyzed using the SWATH DIA method ^1^. The sample analysis order was randomized, and analyses of aliquots of the pooled sample used for library generation were scheduled every 20 runs throughout the sequence as a quality control. MS1 scans were acquired in the Orbitrap with an m/z range of 350 – 1650 m/z at a resolution of 120,000, an AGC target of 4 x 105 and a 3 second cycle time. MS2 scans were then acquired in stepped isolation windows with a 1 Th overlap between each window; first from 350-380 m/z up to 930-960 m/z in increments of 30 Th (i.e. 21 scans with midpoints of 365, 394 … 916, 945), followed by a 100 Th window scan of 959-1059 m/z and a 592 Th scan of 1058-1650 m/z.

Fragmentation of these windows was performed using a normalized collision energy of 25% with a stepped collision energy of 10%. MS2 spectra were acquired in the Orbitrap using a resolution of 30,000, a maximum injection time of 54 ms, an AGC target of 5 x 10^4^ and a scan range of 360-1650 m/z.

**Statistical Analyses**

*Proteomic Data Processing*

Both the DDA fractionated pools and all SWATH samples and pool data were analyzed simultaneously using DIA-NN v1.7.12 ^2^. MS/MS spectra from the DDA pools were searched against UniProt Reference *Homo sapiens* database (retrieved 15/10/2020) with default settings, including 1 missed cleavage, oxidation of methionine as a variable modification and carbamidomethylation of cysteine as a fixed modification, to generate a spectral library. This was then cross referenced against the SWATH data to quantify peptides, and thus proteins, within each SWATH sample and pool run. Precursor FDR was set to 1%.

Subsequent analysis of the DIA-NN output and integration with the clinical data was performed in R v4.0.2. One sample had extremely low average intensity and more than 15% missing values, so was eliminated from the analysis. Across the remaining 185 samples and pools, proteins with more than 50% missing values were eliminated. Intensity values were transformed by VSN (R package ‘vsn’ ^3^), and the remaining missing values were imputed by a k-nearest neighbor approach (R package ‘impute’ ^4^; Supplementary Figure S2).

*Clinical Variable Preprocessing*

Clinical variables were subdivided into DBD-specific variables, DCD-specific variables and jointly applicable variables (see annotation, Supplementary Table ST1). DBD- and DCD-specific variables were considered for the purposes of assessing correlation with outcome but were not considered for statistical analyses applied to the full dataset.

We curated the non-DBD/DCD specific clinical variables to remove those inappropriate for modeling (single-instance categories, variables which duplicated another variable in different units); the final list is given in Supplementary Table ST1. HLA mismatches were modeled as ‘Match Grade’ with levels None; no mismatches, Favorable; no DR and one or fewer B mismatches, and Non-Favorable; at least one DR or two B mismatches (Supplementary Table ST1). Ethnicity was modeled as White vs Other (incorporating Asian, Black, Chinese, Mixed) due to low representation of most ethnicities (encodings as reported by QUOD).

Missing values for eGFR12 were imputed for each sample by finding the ‘set of neighbors’ by 3-month eGFR, i.e. the 10 samples with the closest recipient eGFR at 3 months posttransplant, then taking the mean (ignoring any missing values) within that set of neighbors.

For curated clinical variables, missing values were imputed for each sample in a similar manner. A set of neighbors was built by finding the 10 samples with the smallest Gower’s distance across all variables (multiple data types). We then took a relevant summary statistic of the non-missing values within the set of neighbors for each imputed value. For numeric values, the mean was taken. For categorical values, the most frequent category across the whole dataset was taken, breaking ties by iteratively shrinking the set of neighbors by removing the most distant from the sample to be imputed, until there was a single most frequent category.

*Outcome Association Comparison*

Clinical variable association was calculated depending on the type of each variable in pairwise comparisons. Numeric-numeric comparisons were by squared Pearson’s r (‘R^2^’). Numeric-categorical comparisons were by the proportion of variance explained (eta) from an ANOVA model where the numeric variable was taken as the response. Categorical-categorical comparisons were by Cramér's V.

*Discovery and Evaluation Data Split*

We split our data into discovery and evaluation sets, sampling equally across tertile-stratified eGFR12. Of the 185 samples, 2/3 were used for discovery (‘discovery set’, 118 kidneys), and 1/3 were used for testing (‘evaluation set’, 61 kidneys). Six paired kidneys (3 pairs) were reserved from both sets and used to assess MS and model performance.

*Naïve Feature Selection*

Within the discovery dataset, we modeled outcome against all protein abundance data plus curated clinical variables (Supplementary Table ST2) using Prediction Rule Ensemble (PRE) modeling (R package ‘pre’ ^5^). Briefly, for a given response variable (here, rank-transformed eGFR12), a PRE model identifies a minimal ensemble of prediction functions of the one or more input variables (‘rules’). Functions can include both linear and spline fits as well as decision trees, allowing for detection of nonlinear associations and variable interactions (Figure 1, inset). Each modeling interation includes a LASSO ^6^; as a result when strongly correlated variables are present in a dataset, only one will tend to be selected in the ensemble model for that iteration. Since the PRE algorithm uses a subsampling strategy to generate candidate ensemble rules (see inset, Figure 1), we enforced a fully linear distribution of the response variable by rank-transforming eGFR12. Note that for subsequent linear modeling, we instead used a linearising transform against the UK population distribution (see below) to ensure that model output could be translated back to eGFR12 value in ml/min/1.73 m^2^. PRE was run via the ‘gpe’ function allowing rules to be selected from linear fits, decision tree fits and multivariate adaptive regression spline fits (via R package ‘earth’^7^). Decision tree learning was set to be ‘random forest’-like, with 500 initial trees generated and boosting disabled to allow parallel processing. Other settings (including tree pruning and the parameters for linear and spline fits) were left as default; tree depth was therefore limited to the default 3 decision levels. Ensemble modeling was performed in an iterative manner. At each stage, modeling was performed on the set of predictors that excluded proteins listed in the final ensemble of all previous models. Clinical variables were never excluded. Modeling was repeated for 2000 iterations.

*Linearization of eGFR12 for Regression Modeling*

Estimated Glomerular Filtration Rate (eGFR) values exhibit nonlinear scaling at high eGFR (> ~100 ml/min/1.73 m^2^). To mitigate nonlinear effects while preserving a translation back to standard eGFR scale and ensure data generalized outside our discovery set, we generated a linearized score by transforming measured eGFR12 to be the proportion of eGFR12 values in the NHS Blood and Transplant National Registry (NTxR) between 2016 and 2021 inclusive (6 years) that were less than the measured value (i.e. the quantile of NTxR scores that corresponded to that value). These ‘linearized eGFR12’ values were used for regression modeling; to transform linearized eGFR12 values back to ml/min/1.73 m^2^ scale we used a linear interpolation between the quantiles in NTxR distribution either side of the predicted score. Root-mean-squared error statistics (see below) were calculated on the de-transformed ml/min/1.73 m2 scale for interpretability.

*Regression Modeling*

To model individual protein associations with outcome, we used regularized regression (relaxed LASSO, package ‘glmnet’, using the cv.glmnet function with 20-fold cross validation) to regress each protein, donor age and protein:age terms against linearized eGFR12 (see above) in our discovery dataset, selecting as the final model the model that maximized the shrinkage parameter lambda such that it was within 1 standard error of the lambda value that had the smallest cross-validation error. An independent age-only model was also fit against the discovery dataset using the same parameters. For each protein model and the age-only model we calculated the prediction root-mean-square error (RMSE). Proteins with a higher discovery RMSE than the independent donor age model, or whose model did not retain either the protein or protein:age term were discarded. Final performance of each individual protein model was assessed against the evaluation data, again by RMSE.

*Network Analysis*

The final list of proteins resulting from regression modeling was queried using the Enrichr platform (R package ‘enrichR’ ^8^) against the Reactome 2022 database. Annotation term assignments were filtered at 5% FDR. Interaction networks were generated via STRINGdb ^9^ (R package ‘STRINGdb’)

*Spatial Correlation Analysis*

Processed, normalized Normal, AKI and CKD spatial scRNA-seq expression data generated by Lake *et al.*^10^ were obtained from CELLxGENE (<https://cellxgene.cziscience.com/collections/bcb61471-2a44-4d00-a0af-ff085512674c>). Pseudobulk aggregates of cell counts by cell type and across Normal versus combined AKI and CKI conditions (henceforth ‘Damaged’) were calculated using the AggregateExpression function (R package ‘Seurat’ ^11^). The 539 Proteins identified by LASSO regression filtering were matched to the scRNAseq dataset field “feature_name” by the ‘Genes’ column in the proteomics DIA-NN output. For the full group of 539 proteins, and for sub-groups corresponding to the top 10 Reactome pathways identified by Enrichr analysis, gene-wise fold change across was calculated between Normal and Damaged Cells using the FoldChange function (Seurat) on the scaled pseudobulk expression values. Cell type and full group/pathway sub-group-wise two-tailed t-tests were performed against a null hypothesis of zero fold change across all genes in the full group/sub-group. To correct for multiple testing, q-values were calculated over all comparisons (R package ‘fdrtool’ ^12^), and reported as significant where q<=0.05 (false discovery rate <= 5%).

**SUPPLEMENTARY METHODS REFERENCES**

1.	Gillet LC, Navarro P, Tate S, et al. Targeted data extraction of the MS/MS spectra generated by data-independent acquisition: a new concept for consistent and accurate proteome analysis. *Mol Cell Proteomics*. 2012;11(6):O111.016717.

2.	Demichev V, Messner CB, Vernardis SI, Lilley KS, Ralser M. DIA-NN: neural networks and interference correction enable deep proteome coverage in high throughput. *Nat Methods*. 2020;17(1):41-44.

3.	Huber W, von Heydebreck A, Sultmann H, Poustka A, Vingron M. Variance stabilization applied to microarray data calibration and to the quantification of differential expression. *Bioinformatics*. 2002;18(Suppl 1):S96-S104.

4.	Hastie T, Tibshirani R, Narasimhan B, Chu G. *Impute: Imputation for Microarray Data*.; 2021. http://www.bioconductor.org/packages/release/bioc/html/impute.html

5.	Fokkema M. Fitting prediction rule ensembles with R package pre. *J Stat Softw*. 2020;92(1):1-30.

6.	Friedman JH, Hastie T, Tibshirani R. Regularization Paths for Generalized Linear Models via Coordinate Descent. *J Stat Softw*. 2010;33:1-22.

7.	Milborrow. Derived from mda:mars by T. Hastie and R. Tibshirani. S. *Earth: Multivariate Adaptive Regression Splines*.; 2011. http://CRAN.R-project.org/package=earth

8.	Jawaid W. *EnrichR: Provides an R Interface to “Enrichr.”*; 2021. https://cran.r-project.org/package=enrichR

9.	Szklarczyk D, Kirsch R, Koutrouli M, et al. The STRING database in 2023: protein-protein association networks and functional enrichment analyses for any sequenced genome of interest. *Nucleic Acids Res*. 2023;51(D1):D638-D646.

10.	Lake BB, Menon R, Winfree S, et al. An atlas of healthy and injured cell states and niches in the human kidney. *Nature*. 2023;619(7970):585-594.

11.	Hao Y, Stuart T, Kowalski MH, et al. Dictionary learning for integrative, multimodal and scalable single-cell analysis. *Nat Biotechnol*. 2024;42(2):293-304.

12.	Strimmer K. fdrtool: A versatile R package for estimating local and tail area-based false discovery rates. *Bioinformatics*. 2008;24(12):1461-1462.

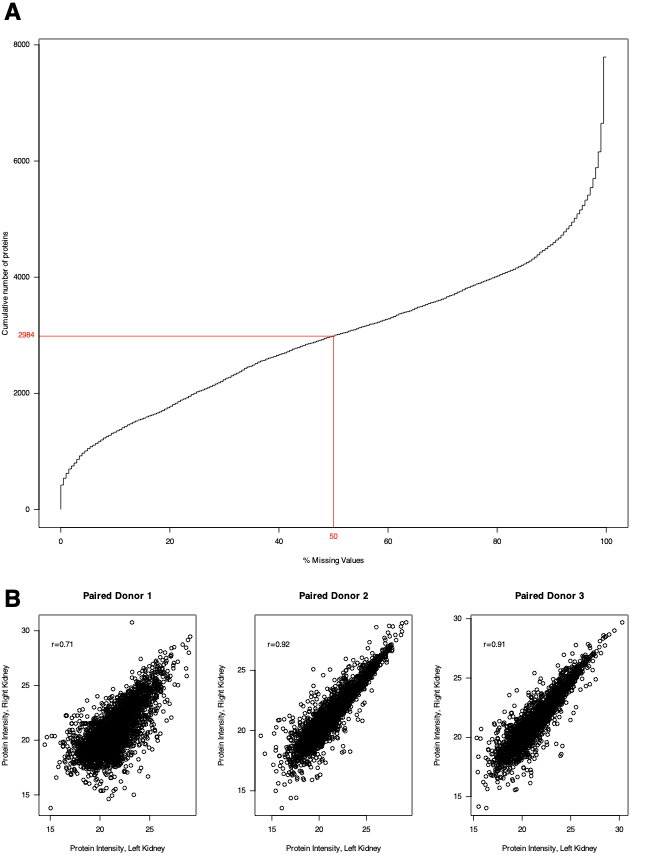
**SUPPLEMENTARY FIGURES**

**Supplementary Figure SF1: Summary of protein quantitation quality**

A: Percentage of missing values across all proteins, showing the cut-off (red line) for poorly quantified proteins excluded from analysis. B: Correlation between quantitation values for each of the 3 sets of paired kidneys.

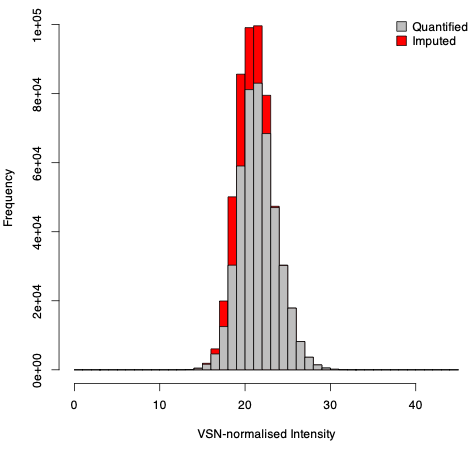
**Supplementary Figure SF2: Summary of protein quantitation imputation**

Histogram showing the fraction of imputed values (red) across distribution of all values.
